# Essential Emergency and Critical Care (EECC) in Ethiopia: A Multicenter mixed-method assessment of Critical Illness Burden, Management, and Implementation Barriers

**DOI:** 10.64898/2026.08.01.26359450

**Authors:** Tesfay Yohannes Ambese, Peniel Kenna Dulla, Fitsum K. Belachew

## Abstract

**Background:** Critical illness is a major contributor to morbidity and mortality in low-resource settings, yet data on its prevalence, location, management, and contextual barriers to care outside intensive care units remain limited in Ethiopia. This study aimed to determine the burden of critical illness, patient outcomes, Essential Emergency and Critical Care (EECC) provision, and barriers to implementation.

**Methods:** This prospective mixed-methods multicenter study was conducted across 12 public hospitals in Ethiopia. A quantitative point prevalence survey assessed all adult inpatients (≥18 years) on a single census day per site using standardized vital sign criteria for critical illness. Patients were followed for 7-day in-hospital mortality, and hospital EECC resource availability was evaluated. A complementary qualitative component consisting of five key informant interviews and one focus group discussion was conducted with healthcare professionals to explore barriers to EECC implementation. Thematic analysis was used for qualitative data.

**Results:** Of 1,077 hospitalized patients, 221 (20.5%) were critically ill. Most critically ill patients (62.0%) were managed in general wards. The 7-day in-hospital mortality was 14.0% among critically ill patients compared to 2.8% in non-critically ill patients. Only 1.8% of critically ill patients received all indicated EECC treatments. Median hospital EECC resource availability was 70.9% (IQR 41.4-79.1%), with no hospital fully equipped across all domains. Qualitative analysis identified seven major themes: challenges in early identification, resource constraints, knowledge gaps, attitudes toward EECC, organizational barriers, staff motivation issues, and delays in timely care delivery. Independent predictors of mortality included male sex, hypertension, cancer, HIV/AIDS, other comorbidities, and presence of critical illness.

**Conclusion:** This critical illness outcome study in Ethiopia demonstrates that one in five hospitalized patients is critically ill, predominantly managed in general wards, with high short-term mortality and profound gaps in basic EECC delivery. Qualitative findings highlight multiple interconnected barriers to effective EECC implementation. Strengthening decentralized EECC through training, resource provision, ward-level protocols, and addressing systemic barriers is urgently needed to improve outcomes in resource-limited settings.

## 1. Introduction

Critical care is an essential component of the healthcare system, providing the necessary health attention in times of life-threatening conditions. The point prevalence of critical illness accounts for 12% of the global population (1,2). In low and low-middle income countries 94% of this care is provided in the health facility; yet almost half of the care is delivered outside of the intensive care unit (ICU) (3). African estimation indicates 69% of critical illnesses are treated in general wards (2).

The COVID-19 pandemic dramatically exposed the vulnerabilities of critical care systems worldwide and underscored the urgent need to strengthen capacity, particularly in resource-limited settings (4). In Ethiopia, pre-pandemic assessments revealed critically low ICU infrastructure, including limited bed capacity, inadequate equipment, and insufficient trained personnel (5). Although a subsequent post-COVID-19 study demonstrated a threefold increase in ICU bed capacity, significant limitations in advanced care, staffing, and overall service readiness continue to persist (6). High bed occupancy further compounds these challenges; a 10% increase in ICU occupancy at the time of admission has been associated with a 7% increase in the odds of mortality (7). Resource strain often results in delays in diagnosis and treatment, reduced individualized care, and worse clinical outcomes (7,8).

Relying exclusively on ICU-based care for critical illness which frequently presents or deteriorates in general wards can lead to delayed interventions, increased morbidity and mortality, overburdened acute care services, staff burnout, and reduced availability of advanced care beds (7,9,10). In recognition of these systemic realities, the Essential Emergency and Critical Care (EECC) initiative promotes the delivery of basic, life-saving interventions at the earliest point of identification, rather than depending solely on scarce ICU resources(11,12). EECC emphasizes simple, low-cost, high-impact actions such as airway management, oxygen therapy, intravenous fluids, and close monitoring that can be effectively implemented in general wards and emergency departments(12–14).

Although various studies have assessed Ethiopia’s critical care capacity by examining ICUs, the true empirical estimation of the prevalence of critical illness and care has yet to be addressed(5,6,15). Understanding this will play a crucial role in shaping critical care provision and help establish frameworks that align with the burden of acute illness. In addition, identifying the location of critically ill patients will aid in pinpointing gaps in care access by decentralizing critical care designs. Given the high cost of ICUS and the fact that most patients require low-cost interventions, understanding the burden and location will inform how cost-effective interventions, such as EECC, can be implemented. This, in turn, can strengthen emergency and outbreak preparedness.

This study aimed to evaluate the burden, location, and management of critical illness, along with short-term patient outcomes and barriers to care. Using a mixed-methods approach, the quantitative component assessed the prevalence of critical illness, provision of EECC, 7-day in-hospital mortality, and predictors of mortality. A complementary qualitative component explored healthcare providers’ perspectives on barriers and facilitators to EECC implementation. This comprehensive assessment seeks to provide actionable evidence to guide context-appropriate improvements in critical care delivery.

## 2. Methodology

### 2.1 Study design and setting

This was a convergent mixed-methods study conducted across 12 public hospitals in Ethiopia (11 level 3 university/referral hospitals and 1 level 2 regional hospital), selected through the Network for Perioperative and Critical Care (N4PCc) collaborating-site network. Data collection took place between June 2025 and September 2025. The study combined a prospective point-prevalence survey of hospitalized patients with qualitative key informant interviews (KIIs) and a focus group discussion (FGD) to assess the burden of critical illness, hospital readiness EECC, and barriers and facilitators to EECC implementation. Quantitative and qualitative strands were collected concurrently and integrated at the interpretation stage through a narrative comparison organized around three shared domains (identification of critical illness, resource availability, and implementation barriers) of findings.

The quantitative component assessed all adult inpatients (aged ≥18 years) present in any ward/department on a single census day per site, chosen by site investigators based on staff availability and logistical feasibility, avoiding weekends and public holidays to capture representative staffing and admission patterns. Psychiatric wards and outpatient areas were excluded. The design was observational, with no intervention performed.

The qualitative component explored healthcare workers’ experiences, perceptions, and practices related to the identification and management of critically ill patients and implementation of EECC within participating hospitals.

### 2.2 Quantitative data collection

Data were collected by trained research healthcare workers at each site, who received standardized training on the study protocol and vital-sign assessment prior to data collection. Investigators measured vital signs (heart rate, oxygen saturation, respiratory rate, systolic blood pressure, and conscious level using the AVPU scale) at the bedside or recorded recent values from clinical notes when direct measurement was not possible. Additional patient data included age, sex, urgency of admission (emergency/acute or elective), main admission category (non-communicable disease, maternal health, trauma, or infection), known chronic conditions or pregnancy, current treatments (oxygen therapy, intravenous fluids, vasopressors, airway interventions), ward type, and ward level.

Hospital-level data on the availability of 67 EECC resources (equipment, consumables, drugs, human resources, training, routines, guidelines, and infrastructure) were collected once per site by using a standardized tool adapted from the EECC assessment framework (11,12).

Patients were followed up at 7 days post-assessment to determine in-hospital status (discharged alive, still in hospital, or died). Patients still hospitalized at day 7 were classified as survivors for the purposes of this analysis. Missing data were excluded on a variable-wise basis; complete-case counts are reported alongside percentages.

### 2.3 Qualitative participant selection and data collection

Purposive sampling was used to recruit healthcare professionals involved in the care of critically ill patients and implementation of EECC. Participants included physicians, nurses, hospital managers, quality improvement officers, emergency and critical care providers, and EECC focal persons. Thirteen healthcare professionals were approached; two declined due to scheduling conflicts and one due to workload, yielding five key informant interviews and one five-person group discussion.

Data were collected through semi-structured KIIs and one FGD, conducted in English by two trained researchers. Interview guides were developed based on the EECC framework, piloted with two senior clinicians at a non-participating hospital to assess clarity and flow, with minor wording revisions made before use, and explored perceptions of critical illness recognition, availability and utilization of EECC resources, knowledge and skills related to EECC, organizational and system-level barriers, staff motivation, and potential strategies for strengthening EECC implementation.

All interviews and the FGD were conducted by trained researchers, audio-recorded with separate informed consent, and supplemented by field notes. Interviews continued until the research team judged thematic saturation had been reached within the scope of this exploratory component.

### 2.4 Outcomes

The co-primary outcomes for the quantitative component were the prevalence of critical illness on the census day and 7-day in-hospital mortality. Critical illness was defined, consistent with prior point-prevalence studies of critical illness (1,2), as the presence of at least one severely deranged vital sign: respiratory rate <8 or >30 breaths/min, oxygen saturation <90%, systolic blood pressure <90 mmHg, heart rate <40 or >130 beats/min, or conscious level responsive to pain or unresponsive on AVPU.

Secondary outcomes included location of care for critically ill patients, provision of EECC treatments (oxygen, intravenous fluids/vasopressors, airway interventions), hospital resource availability, and emergency unit readiness.

For the qualitative component, the outcomes were healthcare workers’ perspectives on barriers and facilitators affecting the identification, management, and delivery of EECC services.

### 2.5 Data analysis

Quantitative data were analyzed descriptively. Continuous variables are presented as medians with interquartile ranges (IQR), and categorical variables as frequencies with percentages. Comparisons between critically ill and non-critically ill patients used χ² tests for categorical data and Mann-Whitney U tests for continuous data. Independent predictors of 7-day mortality were identified using multivariable logistic regression, with covariates selected a priori based on clinical plausibility / via a p<0.10 threshold in univariable analysis, including sex, hypertension, cancer, HIV/AIDS, other comorbidities, and presence of critical illness. All quantitative analyses were performed using Python (pandas, scipy, statsmodels/scikit-learn for regression, and matplotlib for visualization). A p-value <0.05 was considered statistically significant.

Qualitative data were transcribed verbatim and imported into Taguette software for coding and analysis by two coders. Data were analyzed using thematic analysis following Braun and Clarke’s approach. Transcripts were read repeatedly to achieve familiarity with the data, followed by open coding. Codes were grouped into categories and subsequently organized into broader themes through an iterative process involving discussion among the research team, with disagreements resolved by consensus. Emerging themes were reviewed, refined, and agreed upon by consensus. Representative quotations were selected to illustrate key findings.

### 2.6 Ethical considerations

Ethical approval was obtained from the Institutional Review Board (IRB) of the Armauer Hansen Research Institute (AHRI) (protocol number PQ49/23). For the quantitative component, the requirement for individual written informed consent was waived by the IRB given the observational nature of the study; data were anonymized to ensure confidentiality. For the qualitative component, all KII and FGD participants provided separate verbal informed consent prior to participation, including consent to audio recording. The Ministry of Health (MOH), a key collaborator in the project, facilitated communication with hospitals through official letters and ensured their cooperation during data collection.

## 3. Results

### 3.1 Patient-level outcomes

A total of 1,077 patients were enrolled, of whom 221 (20.5%) were identified as critically ill. The median age was 35 years [IQR 25–50], with critically ill patients slightly older (36 [27–55] years) than those without critical illness (34 [25–50] years). Sex distribution was similar across groups, with 48.9% female and 51.1% male. Chronic conditions were reported in both groups, with higher proportions of COPD/asthma (16.8%), heart disease (12.7%), and tuberculosis (11.8%) among critically ill patients. Overall, 25.0% of critically ill patients reported no chronic illness compared to 48.7% of non-critically ill patients. Most admissions were emergencies (83.0% overall; 95.5% among critically ill patients). In terms of diagnostic categories, non-communicable diseases (42.2%) and trauma (23.9%) were most common overall, while non-communicable diseases (48.4%) and infections (27.6%) predominated among critically ill patients.

Ward distribution showed that most patients were admitted to general wards (82.6%), though critically ill patients were more frequently managed in medical wards (62.0%), high-dependency units (18.1%), or ICUs (19.9%). Physiological parameters differed between groups: critically ill patients had higher median heart rate (105 vs. 86 bpm) and respiratory rate (28 vs. 21 breaths/min), lower oxygen saturation (92% vs. 96%), and lower systolic blood pressure (108 vs. 115 mmHg). Airway compromise was more common among critically ill patients, with 24.4% showing partial obstruction compared to 2.1% in non-critically ill patients. Consciousness level also varied, with 65.2% of critically ill patients fully alert compared to 95.7% of non-critically ill patients.

**Table 1:**
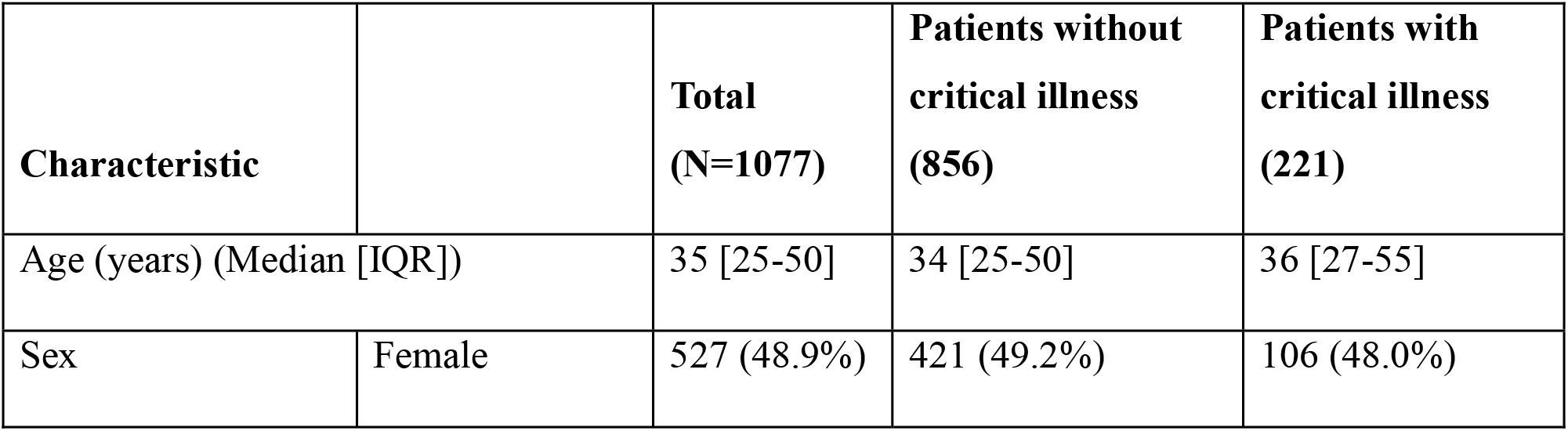
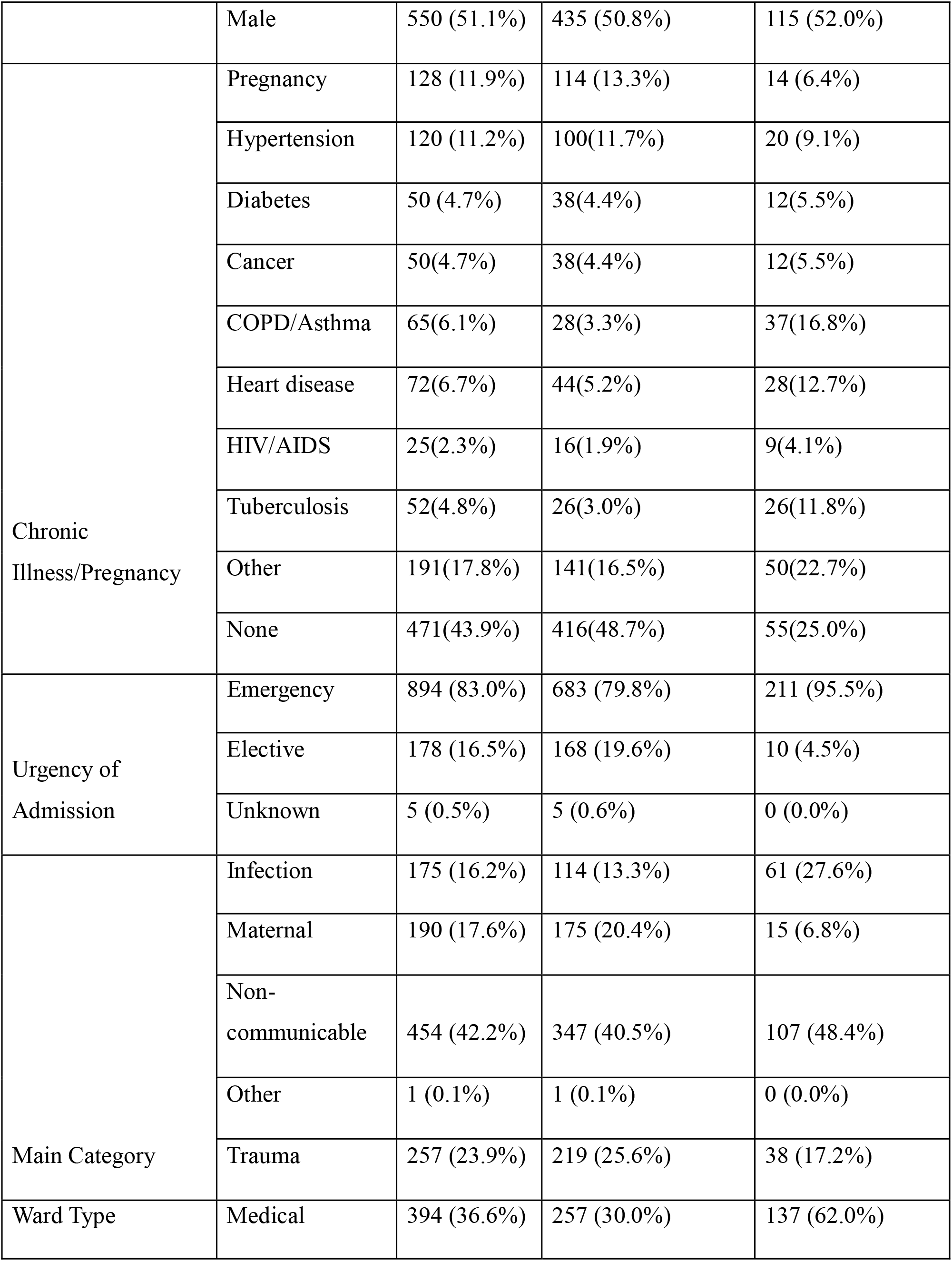
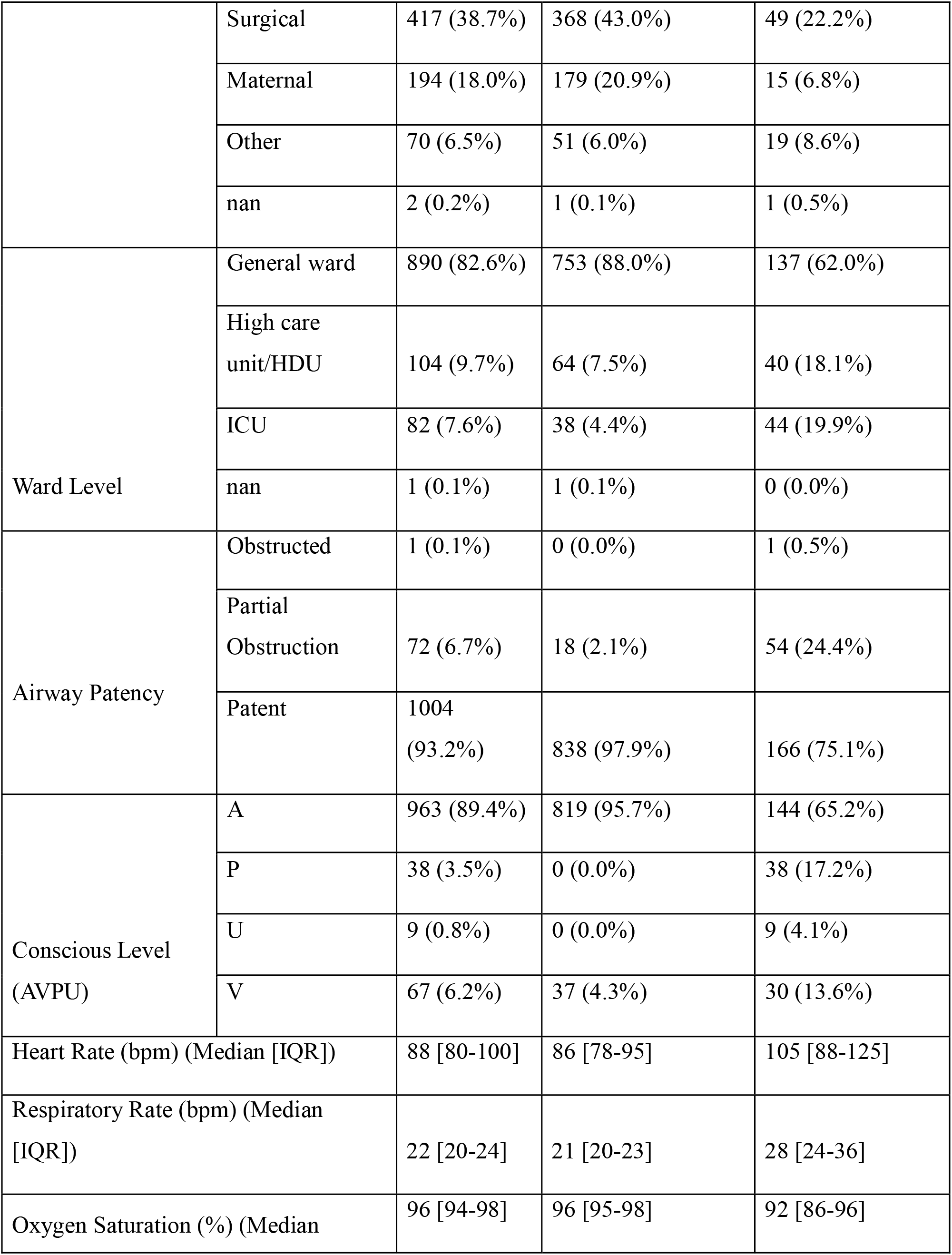
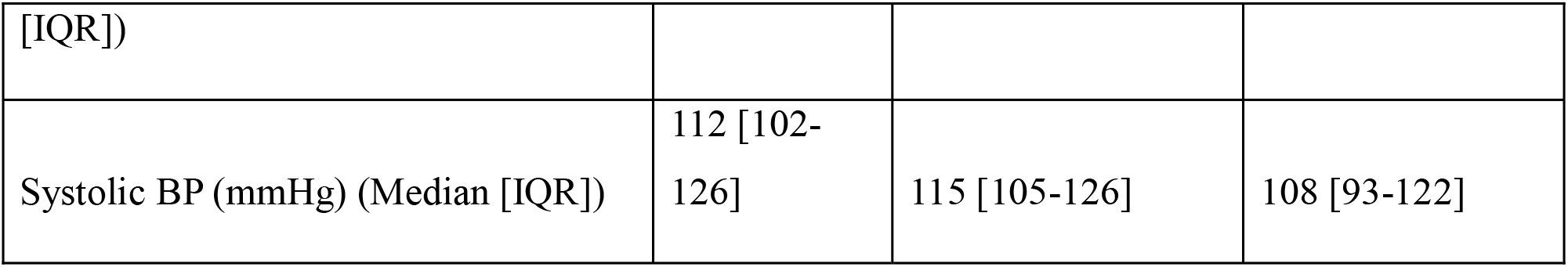
Baseline characteristics.

As shown in Table 2, the point prevalence of critical illness among 215 hospitalized patients was 20.5% (221/1077) of 47 (4.4%) were classified based on impaired consciousness, 86 (8.0%) by circulatory compromise, and 149 (13.8%) by respiratory dysfunction. Most critically ill patients met a single criterion (165; 15.3%), while 51 (4.7%) fulfilled two criteria and 5 (0.5%) met all three. The overall 7-day in-hospital mortality was 55 (5.1%). Mortality was substantially higher among critically ill patients (31 deaths; 14.0%) compared to non-critically ill patients (24 deaths; 2.8%).

**Table 2:** Point prevalence of critical illness and mortality.

| Characteristics | N (%) |
| --- | --- |
| Prevalence of critical illness |  |
| Critically ill patients | 221 (20.5%) |
| **Critical illness categories** |  |
| Patients defined as critically ill by conscious level | 47 (4.4%) |
| Patients defined as critically ill by circulatory | 86 (8.0%) |
| Patients defined as critically ill by respiratory | 149 (13.8%) |
| Patients defined as critically ill by one criterion only | 165 (15.3%) |
| Patients defined as critically ill by two criteria | 51 (4.7%) |
| Patients defined as critically ill by three criteria | 5 (0.5%) |
| **7 day in-hospital mortality** |  |
| Mortality (whole cohort) | 55(5.1%) |
| Mortality (critically ill patients) | 31(14.0%) |
| Mortality (non-critically ill patients) | 24(2.8%) |

Figure 1 shows the distribution of seven-day in-hospital mortality according to location of care. Overall mortality was 5.1% (55/1077). Mortality in the general ward was 3.1% (28/889), in the high-dependency unit (HDU) 10.6% (11 of 104), and in the intensive care unit (ICU) 19.5% (16/82).

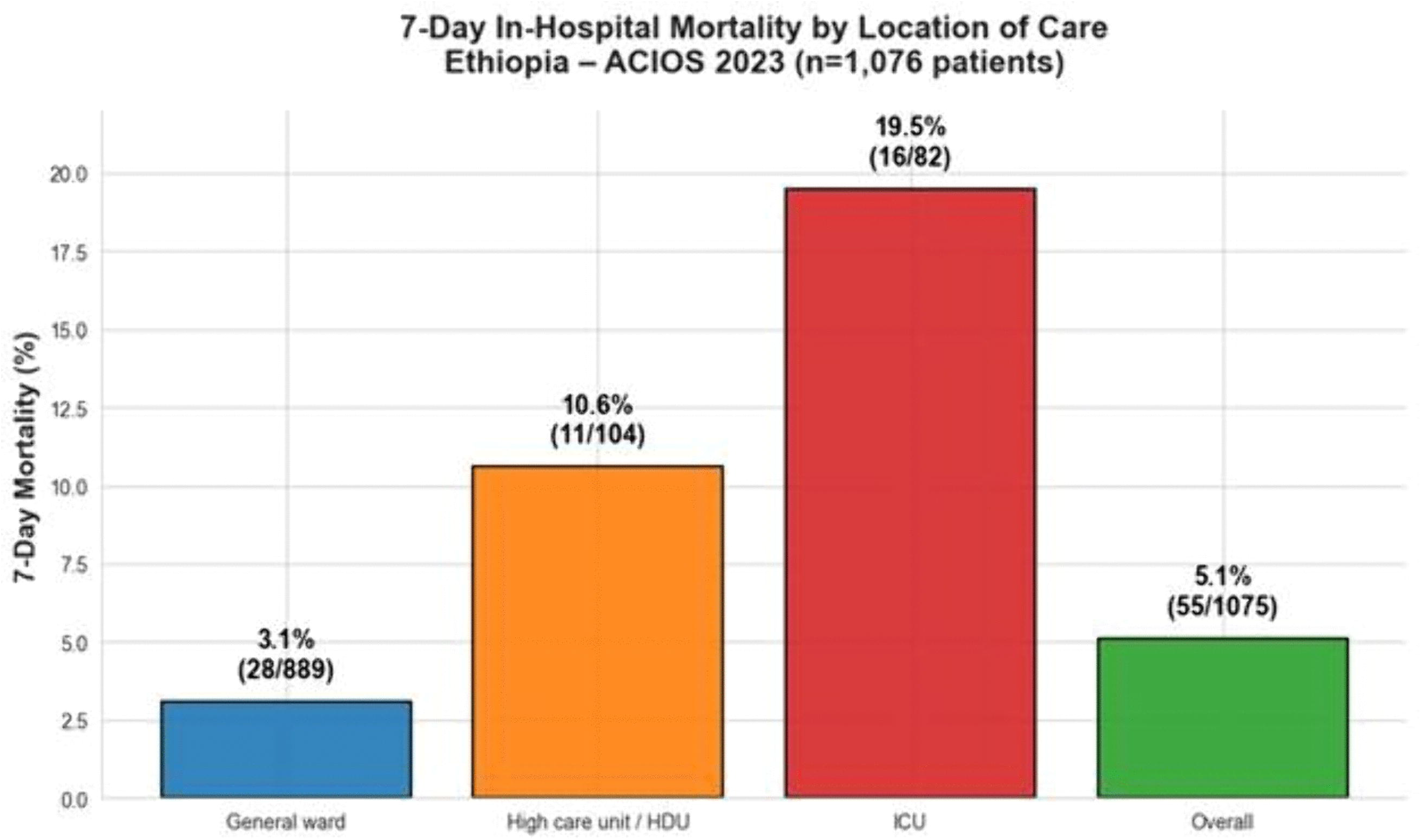

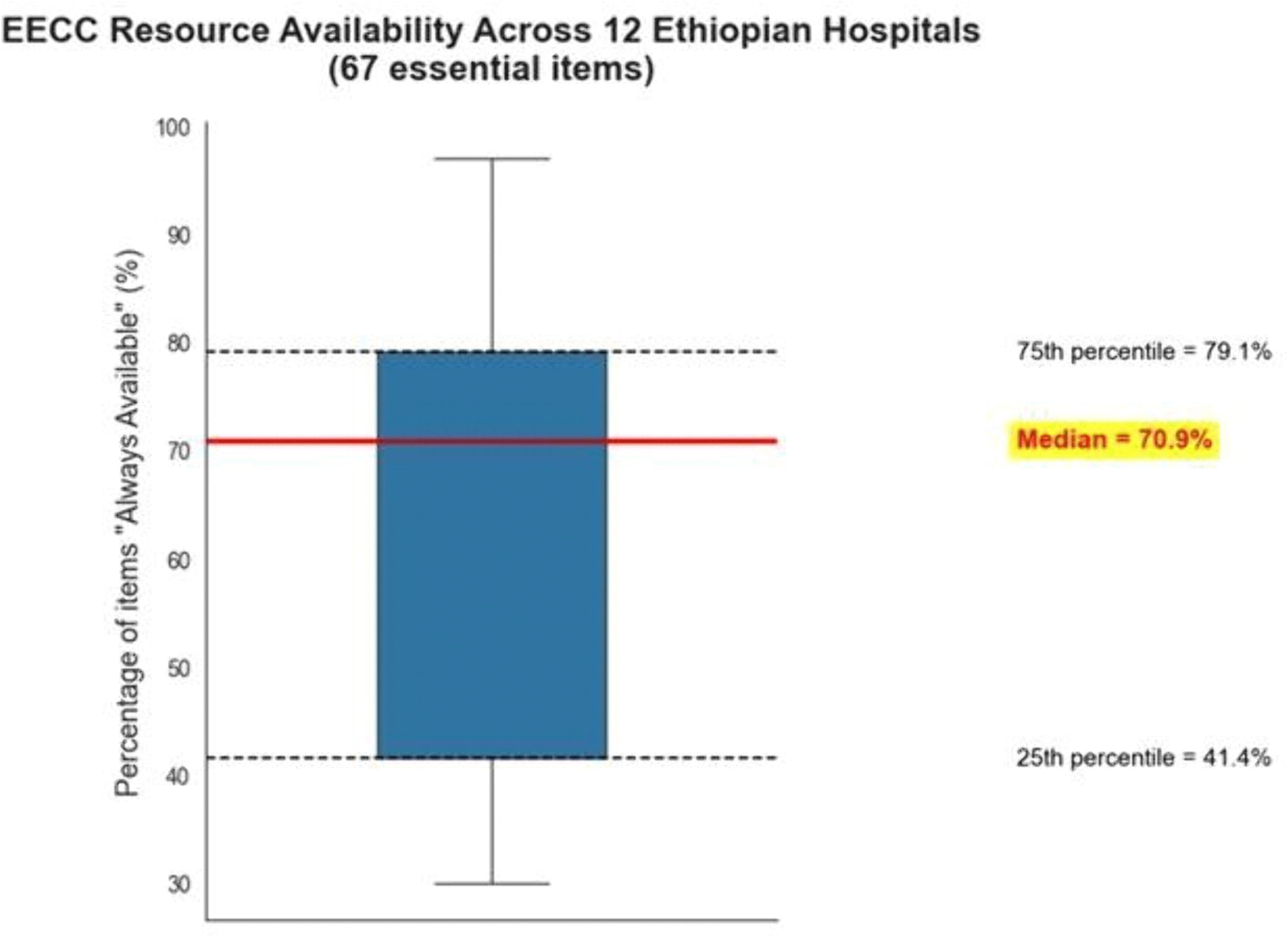

As shown in Table 3, among the 221 critically ill patients, treatment data were available for 220, and only 4 patients (1.8%) received all indicated EECC treatments, while 216 (98.2%) received partial or none. Of those defined by respiratory criteria (n=149), 107 (71.8%) received oxygen. Among patients defined by circulatory criteria (n=86), 53 (61.6%) received intravenous fluids, 24 (27.9%) received vasopressors, and 54 (62.8%) received either intravenous fluids or vasopressors. For patients defined by conscious level criteria (n=46), 31 (67.4%) received an airway intervention, 6 (13.0%) were placed in the recovery position, and 36 (78.3%) received either an airway intervention or recovery positioning.

**Table 3:**
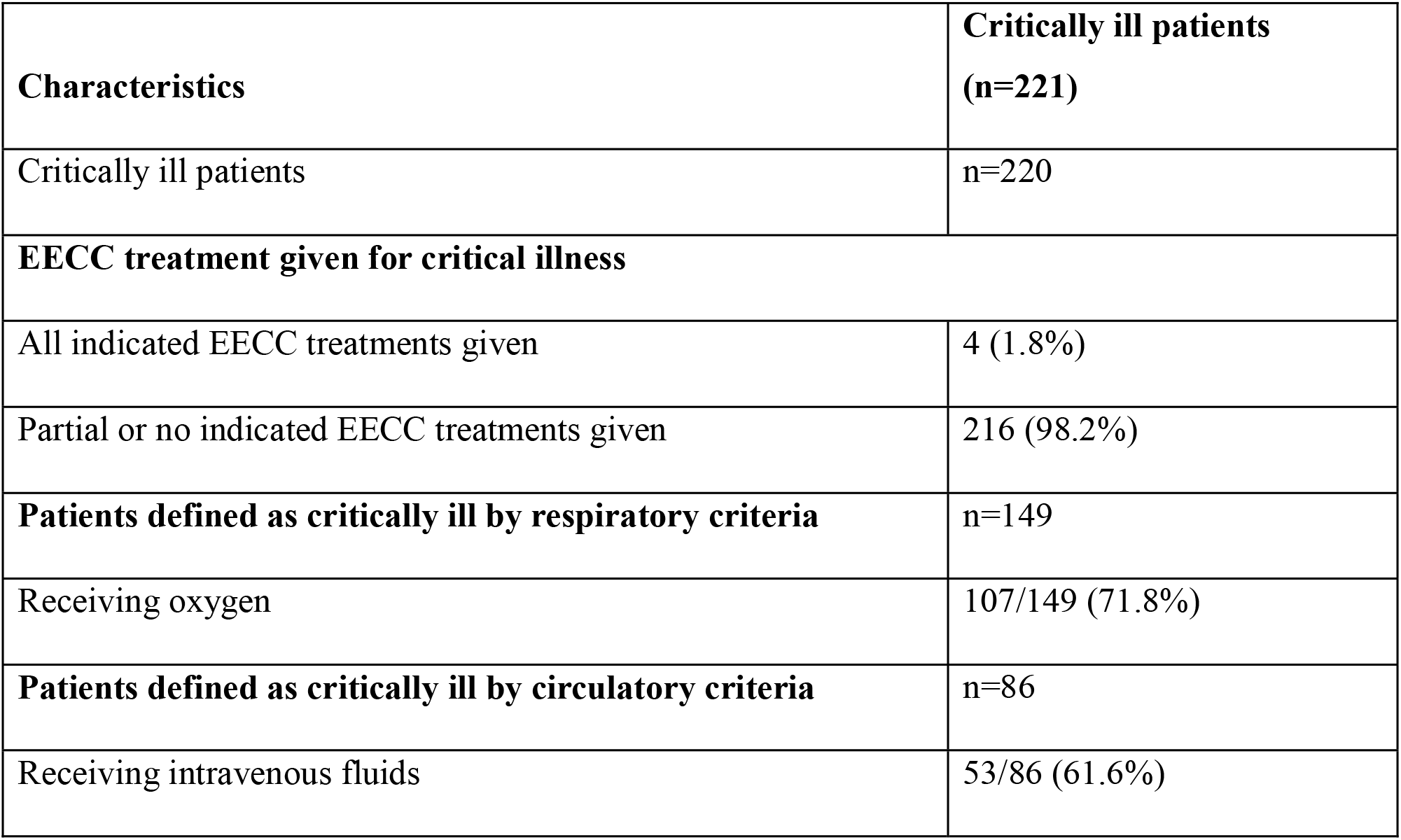
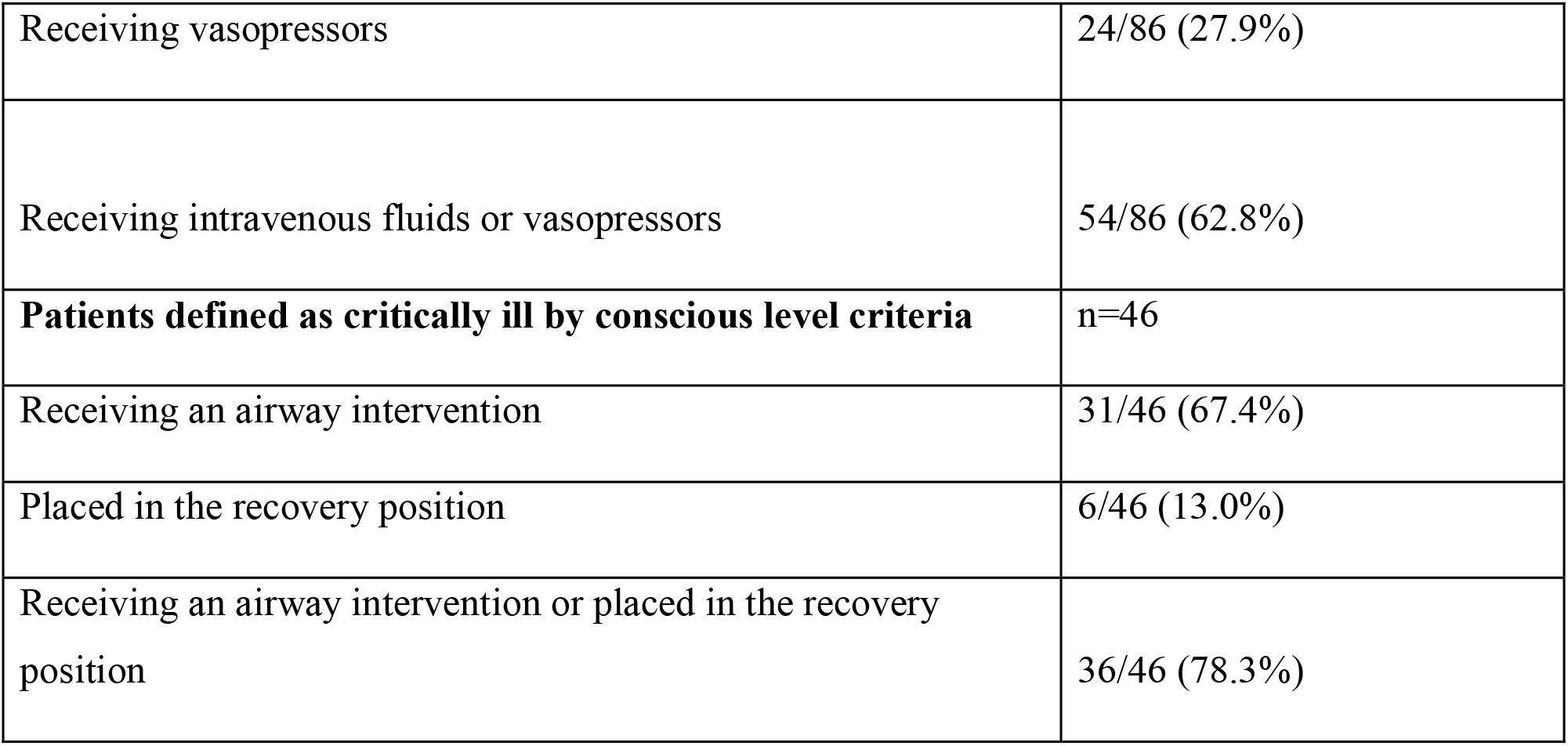
The EECC treatments given to critically ill patients.

| Characteristics | Critically ill patients (n=221) |
| --- | --- |
| Critically ill patients | n=220 |
| **EECC treatment given for critical illness** |  |
| All indicated EECC treatments given | 4 (1.8%) |
| Partial or no indicated EECC treatments given | 216 (98.2%) |
| **Patients defined as critically ill by respiratory criteria** | n=149 |
| Receiving oxygen | 107/149 (71.8%) |
| **Patients defined as critically ill by circulatory criteria** | n=86 |
| Receiving intravenous fluids | 53/86 (61.6%) |
| Receiving vasopressors | 24/86 (27.9%) |
| Receiving intravenous fluids or vasopressors | 54/86 (62.8%) |
| **Patients defined as critically ill by conscious level criteria** | n=46 |
| Receiving an airway intervention | 31/46 (67.4%) |
| Placed in the recovery position | 6/46 (13.0%) |
| Receiving an airway intervention or placed in the recovery position | 36/46 (78.3%) |

In the multivariable logistic regression analysis assessing predictors of 7-day in-hospital mortality (Table 4), several variables were found to be significantly associated with increased odds of death. Male patients had more than twice the odds of mortality compared to females (AOR = 2.24, 95% CI: 1.09-4.61, *p* = 0.028). Hypertension was independently associated with higher mortality (AOR = 2.93, 95% CI: 1.25-6.89, *p* = 0.014), while patients with cancer had nearly six-fold increased odds of death (AOR = 5.92, 95% CI: 2.23-15.71, *p* < 0.001). Similarly, HIV/AIDS was associated with a four-fold increase in mortality risk (AOR = 4.32, 95% CI: 1.09-17.06, *p* = 0.037). The presence of other comorbid conditions also significantly increased mortality risk (AOR = 3.65, 95% CI: 1.93-6.89, *p* < 0.001).

**Table 4:** Multiple logistic regression for 7-day in-hospital mortality.

| Variables |  | AOR (95% CI) | p-value |
| --- | --- | --- | --- |
| Age |  | 1.01 (0.99, 1.03) | 0.518 |
| Sex | Female | Reference |  |
|  | Male | 2.24 (1.09, 4.61) | 0.028* |
| Pregnant |  | 0.57 (0.08, 3.99) | 0.572 |
| Hypertension |  | 2.93 (1.25, 6.89) | 0.014* |
| Diabetes |  | 0.36 (0.07, 1.80) | 0.212 |
| Cancer |  | 5.92 (2.23, 15.71) | <0.001* |
| COPD/Asthma |  | 1.22 (0.37, 4.00) | 0.748 |
| Heart Disease |  | 0.91 (0.32, 2.60) | 0.867 |
| HIV/AIDS |  | 4.32 (1.09, 17.06) | 0.037* |
| Tuberculosis |  | 0.92 (0.25, 3.47) | 0.908 |
| Others |  | 3.65 (1.93, 6.89) | <0.001* |
| Urgency admission | Elective | Reference |  |
|  | Emergency | 1.71 (0.56, 5.17) | 0.346 |
| Main category for admission | NCD |  |  |
|  | Maternal | 1.35 (0.29, 6.23) | 0.697 |
|  | Trauma | 0.41 (0.14, 1.16) | 0.093 |
|  | Infection | 0.82 (0.34, 1.94) | 0.647 |
| Critically ill | No | Reference |  |
|  | Yes | 5.18 (2.75, 9.73) | <0.001* |
| NCD: Non-communicable disease; COPD=chronic obstructive pulmonary disease. |  |  |  |

Critically ill patients had markedly higher odds of death compared to non-critical admissions (AOR = 5.18, 95% CI: 2.75-9.73, *p* < 0.001). In contrast, age, pregnancy status, diabetes, COPD/asthma, heart disease, tuberculosis, urgency of admission, and admission category (maternal, trauma, or infection) were not significantly associated with 7-day in-hospital mortality in the adjusted model.

### 3.2 Hospital resource availability

Across the 12 participating hospitals, the median availability of resources for EECC was 47.5 out of 67 items (70.9%), with an interquartile range of 41.4% to 79.1%. Availability varied across domains: human resources were most commonly (7/12 hospitals; 58.3%), followed by training (41.7%), equipment (25.0%), guidelines (25.0%), and infrastructure (25.0%). Full availability of drugs was reported in 2 hospitals (16.7%), while none had complete access to consumables. No hospital had all domains fully equipped for EECC.

**Table 5:**
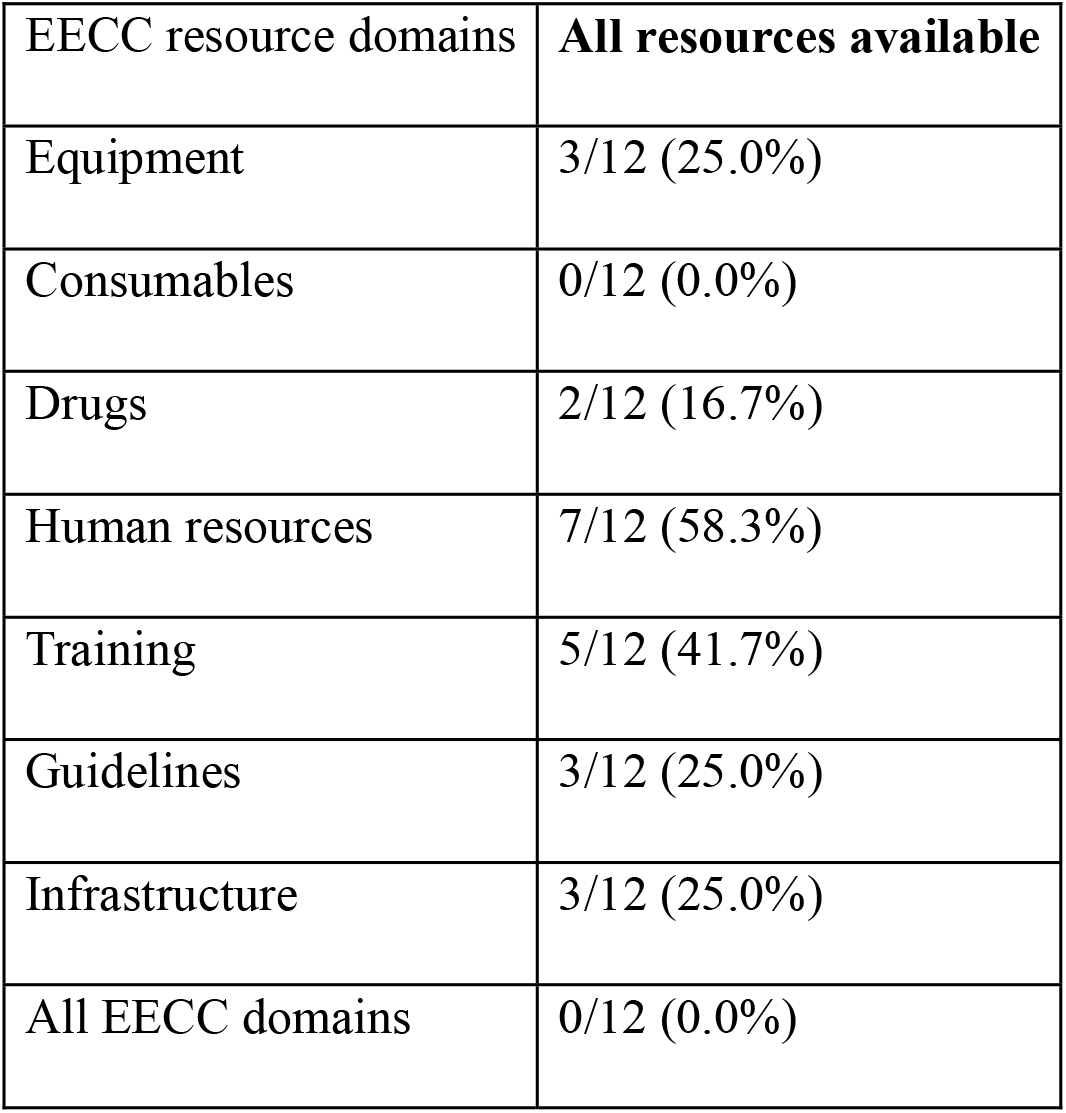
Summary of domain resources available for Essential Emergency and Critical Care (EECC)

| EECC resource domains | All resources available |
| --- | --- |
| Equipment | 3/12 (25.0%) |
| Consumables | 0/12 (0.0%) |
| Drugs | 2/12 (16.7%) |
| Human resources | 7/12 (58.3%) |
| Training | 5/12 (41.7%) |
| Guidelines | 3/12 (25.0%) |
| Infrastructure | 3/12 (25.0%) |
| All EECC domains | 0/12 (0.0%) |

As shown in Figure 2, the availability of EECC resources across the 12 participating hospitals demonstrated substantial variability. The median percentage of the 67 essential EECC items available was 70.9%, with an interquartile range (IQR) of 41.4% to 79.1%. Per-hospital availability ranged widely from 29.9% to 97.0%.

### 3.3 Qualitative Findings

A total of 6 key informant interviews and 1 focus group discussions were conducted among healthcare professionals involved in the delivery of emergency and critical care services. Participants included physicians, nurses, hospital leaders, and quality improvement personnel. Thematic analysis identified seven major themes influencing the implementation of Essential Emergency and Critical Care (EECC): (1) challenges in early identification of critical illness, (2) resource and readiness constraints, (3) knowledge and competency gaps, (4) attitudes and perceptions toward EECC, (5) organizational and system-level barriers, (6) motivation and staff engagement challenges, and (7) barriers to timely delivery of EECC.

#### Theme 1: Challenges in Early Identification of Critical Illness

Participants reported that delayed recognition of critically ill patients was a major challenge. High patient volumes, inadequate staffing, poor monitoring practices, and limited interpretation of abnormal vital signs contributed to missed opportunities for early intervention. Participants also described communication and escalation delays, particularly within the hierarchical structure of teaching hospitals.

Several participants highlighted that although vital signs were often measured, abnormal findings were not consistently interpreted or acted upon, resulting in delayed identification of deterioration.

#### Theme 2: Resource and Readiness Constraints

Lack of essential equipment, medications, and consumable supplies emerged as one of the most frequently reported barriers. Participants described shortages of pulse oximeters, oxygen delivery devices, suction machines, perfusers, and emergency medications. In addition to shortages, participants noted challenges related to equipment maintenance, accessibility, and procurement delays.

Resource distribution was reported to vary across hospital units, with intensive care and high-dependency units generally better equipped than general wards and outpatient departments. Infrastructure limitations, including lack of dedicated spaces for critically ill patients, further constrained EECC implementation.

#### Theme 3: Knowledge and Competency Gaps

Participants reported variability in EECC knowledge and practical skills among healthcare workers. Limited training coverage, inadequate refresher training, and insufficient simulation-based learning opportunities were identified as key factors contributing to competency gaps.

Although EECC training initiatives had been introduced, participants indicated that training often relied on didactic lectures and reached only a fraction of healthcare workers. Lack of routine competency assessment and feedback mechanisms were also reported.

#### Theme 4: Attitudes and Perceptions Toward EECC

A recurring finding was the perception that advanced critical care interventions received greater attention than basic life-saving care. Participants explained that healthcare workers often focused on transferring patients to intensive care units or seeking advanced interventions before initiating essential supportive measures.

Many participants emphasized that EECC was sometimes viewed as less important than definitive or specialized care, resulting in delays in the initiation of simple but potentially life-saving interventions.

#### Theme 5: Organizational and System-Level Barriers

Participants identified several health system factors that influenced EECC implementation. These included staffing shortages, budget limitations, procurement inefficiencies, and inadequate integration of early warning systems into electronic medical records.

While participants generally perceived hospital leadership as supportive of EECC implementation, they noted that broader system-level constraints limited the ability of hospitals to address staffing and resource gaps.

#### Theme 6: Motivation and Staff Engagement Challenges

Healthcare workers described several factors affecting motivation to provide EECC. These included lack of incentives, limited recognition for good performance, emotional exhaustion associated with caring for critically ill patients, and insufficient staff engagement in quality improvement initiatives.

Participants emphasized the importance of acknowledging healthcare workers’ contributions and creating opportunities for active participation in EECC implementation efforts.

#### Theme 7: Barriers to Timely Delivery of EECC

Participants reported that delays in providing EECC were multifactorial. Workload pressures, resource shortages, delayed escalation of care, and patient-related financial barriers all contributed to delays in treatment initiation.

Challenges were reported to be particularly pronounced during weekends, holidays, and night shifts when staffing levels were reduced. Participants also noted that inability of patients to purchase essential medications or supplies could delay treatment even when healthcare workers recognized the need for urgent intervention.

## 4. Discussion

This multicenter prospective mixed method point-prevalence study provides one of the first comprehensive evaluations of the burden, location, management, and short-term outcomes of critical illness in Ethiopian public hospitals. It reveals that one in five (20.5%) hospitalized adult patients fulfilled objective vital sign-based criteria for critical illness. Notably, the majority (62%) of these patients were managed in general wards rather than dedicated high-dependency units (HDUs) or intensive care units (ICUs). The 7-day in-hospital mortality rate among critically ill patients reached 14.0%, more than five times higher than the 2.8% observed among non-critically ill patients. Delivery of EECC interventions was alarmingly inadequate, with only 1.8% of critically ill patients receiving the full package of indicated treatments. Hospital-level EECC resource availability was incomplete (median 70.9% of 67 items), and no participating facility demonstrated full capacity across all assessed domains. These findings expose critical gaps in the early identification and timely management of life-threatening conditions in resource-constrained settings and strongly advocate for the urgent scaling of decentralized EECC approaches.

The observed 20.5%-point prevalence of critical illness exceeds the commonly cited global estimate of approximately 12% and aligns closely with data from the African Critical Illness Outcomes Study (ACIOS), which documented substantial inter-country variation across 22 African nations(2). Our results confirm that critical illness represents a major and under-appreciated burden within Ethiopian hospitals. Crucially, most critically ill patients were cared for in general wards, consistent with the continental estimate that 69% of critical care is delivered outside ICUs(2). This pattern underscores a fundamental limitation of ICU-centric models in LMICs, where critical illness frequently manifests or deteriorates in non-specialized wards due to delayed recognition and resource constraints(3).

The 14% 7-day mortality among critically ill patients, although somewhat lower than the ∼21% reported in the ACIOS, remains unacceptably high and highlights persistent challenges in critical care delivery. Mortality rates increased with the level of care (3.1% in general wards, 10.6% in HDUs, and 19.5% in ICUs), which likely reflects selection bias toward higher severity cases in advanced units(2). Multivariable analysis identified male sex, hypertension, cancer, HIV/AIDS, and other comorbidities as independent predictors of mortality, alongside the presence of critical illness itself. These associations mirror patterns described in other sub-Saharan African studies, where late presentation, comorbid burden, and delays in basic interventions significantly worsen prognosis (7,13). Although post-COVID-19 initiatives have achieved a threefold expansion in ICU bed capacity in Ethiopia, our data demonstrate that limitations in advanced care infrastructure and, more importantly, deficiencies in foundational EECC continue to drive preventable mortality(6).

A particularly concerning finding was the profound mismatch between the identification of critical illness and the actual provision of lifesaving EECC interventions. Despite reasonable availability of certain resources (e.g., oxygen supply and basic human resources), only 1.8% of critically ill patients received all indicated treatments. Oxygen therapy was administered to 71.8% of patients with respiratory criteria, circulatory support to 62.8% of those with shock, and airway interventions to 67.4% with altered consciousness. These gaps in basic, low-cost interventions directly contribute to the observed mortality and are consistent with previous national assessments that highlighted deficiencies in equipment, consumables, training, and standardized protocols(5,15). The wide variability in EECC resource availability across the 12 hospitals (29.9%-97.0%) further illustrates systemic inequities that undermine equitable critical care delivery.

The qualitative findings from key informant interviews and focus group discussions provide important context for these quantitative gaps. Seven major themes emerged as key barriers to effective EECC implementation: challenges in early identification of critical illness, resource and readiness constraints, knowledge and competency gaps, attitudes and perceptions toward EECC, organizational and system-level barriers, motivation and staff engagement challenges, and barriers to timely delivery of EECC. These themes highlight that suboptimal EECC delivery is not only due to resource shortages but also stems from systemic, behavioral, and organizational factors such as delayed recognition of deterioration, preference for ICU transfer over basic interventions, and low staff motivation. Together, the mixed methods results demonstrate that addressing these interconnected barriers is essential for translating resource availability into effective bedside care (11,12).

## Strengths and limitations

This study has several strengths, including its prospective multicenter design across 12 diverse public hospitals, standardized vital sign-based definition of critical illness (aligned with ACIOS and EECC frameworks), bedside data collection, and 7-day outcome follow-up. The integration of patient-level clinical data with hospital-level resource assessments provides a multidimensional view of critical care challenges in Ethiopia. Limitations include the single-day census design, which may introduce temporal or seasonal bias; the lack of longer-term outcomes or detailed complication tracking; reliance on self-reported resource availability, which may overestimate functionality; and restriction to public hospitals, potentially limiting generalizability to private facilities or more remote rural settings. Despite these constraints, the findings offer robust, actionable insights relevant to anaesthesia and critical care practice in similar LMIC contexts.

### Implications for practice and policy

This critical illness outcome study in Ethiopia clearly demonstrates that critical illness is common, predominantly managed in general wards, and associated with high short-term mortality and major shortfalls in basic care. For anesthesiologists and critical care providers, these results highlight the need to extend expertise beyond traditional ICU boundaries through training in EECC principles, development of ward-based early warning systems, and establishment of rapid response protocols(7,11). At the policy level, national health authorities and international partners should prioritize the integration of EECC into existing emergencies, ward, and surgical care pathways. Investments in low-cost, high-yield interventions such as improved oxygen delivery systems, consumables, staff training, and documentation tools offer a cost-effective route to reducing preventable deaths and strengthening resilience against future outbreaks or surges in critical illness(3,12,16).

By addressing these gaps, Ethiopia and other resource-limited settings can move toward a more resilient, decentralized model of critical care that aligns with the realities of patient presentation and available resources, ultimately improving outcomes for the most vulnerable hospitalized patients.

## 5. Conclusion

This multicenter prospective mixed-methods point prevalence study demonstrates that critical illness is highly prevalent in Ethiopian public hospitals, affecting one in five (20.5%) hospitalized adult patients. Most of critically ill patients are managed in general wards, where delivery of basic life-saving EECC interventions remains critically inadequate. The 7-day in-hospital mortality of 14.0% among critically ill patients, combined with profound gaps in EECC provision and incomplete hospital resource availability, highlight major deficiencies in the current system of critical care delivery. Qualitative findings further reveal multiple interconnected barriers, including challenges in early identification, resource constraints, knowledge gaps, unfavorable attitudes toward basic care, and systemic obstacles that hinder timely EECC implementation.

These findings underscore that reliance on ICU-centric models is insufficient to address the burden of critical illness in resource-limited settings. Strengthening decentralized EECC through targeted training of frontline healthcare workers, reliable provision of low-cost essential resources, implementation of ward-based early warning and response protocols, and addressing behavioral and organizational barriers offers a feasible, high-impact, and cost-effective strategy to reduce preventable mortality.

National health authorities, hospital administrators, and international partners should prioritize the integration of EECC into routine emergency, ward, and surgical care pathways as a core component of health system strengthening in Ethiopia. Addressing these gaps will not only improve clinical outcomes for critically ill patients but will also enhance overall emergency preparedness and resilience against future disease outbreaks and surges in critical illness across similar low-resource African settings.

## Declarations

### Author Contributions

**Tesfay Yohannes Ambese:** Conceptualization, Methodology, Formal analysis, Data curation, Writing – original draft, Writing – review & editing.

**Fitsum K. Belachew:** Conceptualization, Supervision, Writing – review & editing.

**Peniel Kenna Dulla:** Investigation, Data curation, Writing – review & editing. All authors read and approved the final manuscript.

### Informed Consent

As this was a prospective observational point prevalence study with no intervention, a waiver of individual informed consent was granted by the IRB. Data were collected as part of routine clinical care assessment, and all patient information was anonymized to ensure confidentiality.

## Data Availability

The datasets generated and/or analyzed during the current study are available from the corresponding author on reasonable request. De-identified participant data and analysis code can be shared upon approval by the institutional ethics committee.

