## Supplementary file for "Essential Emergency and Critical Care (EECC) in Ethiopia: A Multicenter mixed-method assessment of Critical Illness Burden, Management, and Implementation Barriers"

**Appendix**

**Supplementary Table 1**: Resources available for the provision of Essential Emergency and Critical Care (EECC)

| **Equipment** | **Always** | **Sometimes** | **Never** | **Do not know** |
| --- | --- | --- | --- | --- |
| Clock with second hand | 3/12 (25%) | 8/12 (66.6%) | 1/12 (8.3%) | 0/12 (0%) |
| Pulse oximeter and probe | 11/12 (91.6%) | 0/12 (0%) | 0/12 (0%) | 0/12 (0%) |
| Blood pressure monitoring equipment | 11/12 (91.6%) | 1/12 (8.3%) | 0/12 (0%) | 0/12 (0%) |
| Blood pressure cuffs of different sizes | 6/12 (50%) | 6/12 (50%) | 0/12 (0%) | 0/12 (0%) |
| Light source | 6/12 (50%) | 5/12 (41.6%) | 1/12 (8.3%) | 0/12 (0%) |
| Thermometer | 9/12 (75%) | 3/12 (25%) | 0/12 (0%) | 0/12 (0%) |
| Suction machine | 10/12 (83.3%) | 2/12 (16.6%) | 0/12 (0%) | 0/12 (0%) |
| Oxygen supply 24h/day | 11/12 (91.6%) | 1/12 (8.3%) | 0/12 (0%) | 0/12 (0%) |
| Flow meter | 12/12 (100%) | 0/12 (0%) | 0/12 (0%) | 0/12 (0%) |
| Leak-free connectors from oxygen source to tubing | 9/12 (75%) | 1/12 (8.3%) | 1/12 (8.3%) | 1/1 (8.3%) |
| Bag valve mask | 7/12 (58.3%) | 5/12 (41.6%) | 0/12 (0%) | 0/12 (0%) |
| Sharps disposal container | 10/12 (83.3%) | 1/12 (8.3%) | 0/12 (0%) | 1/12 (8.3%) |
| External heat source | 3/12 (25%) | 8/12 (66.6%) | 1/12 (8.3%) | 0/12 (0%) |
| **Consumables** |  |  |  |  |
| Soap or hand disinfectant | 5/12 (41.6%) | 7/12 (58.3%) | 0/12 (0%) | 0/12 (0%) |
| Examination gloves | 8/12 (66.6%) | 4/12 (33.3%) | 0/12 (0%) | 0/12 (0%) |
| Suction catheters of paediatric and adult sizes | 3/12 (25%) | 9/12 (75%) | 0/12 (0%) | 0/12 (0%) |
| Guedel airways of paediatric and adult sizes | 5/12 (41.6%) | 6/12 (50%) | 1/12 (8.3%) | 0/12 (0%) |
| Pillows | 8/12 (66.6%) | 4/12 (33.3%) | 1/12 (8.3%) | 0/12 (0%) |
| Oxygen tubing | 11/12 (91.6%) | 1/12 (8.3%) | 0/12 (0%) | 0/12 (0%) |
| Oxygen nasal prongs | 10/12 (83.3%) | 2/12(16.6%) | 0/12 (0%) | 0/12 (0%) |
| Oxygen face masks of pediatric and adult sizes | 9/12 (75%) | 4/12 (33.3%) | 0/12 (0%) | 0/12 (0%) |
| Oxygen face masks with reservoir bags of pediatric and adult sizes | 4/12 (33.3%) | 8/12 (66.6%) | 0/12 (0%) | 0/12 (0%) |
| Masks for Bag Valve Mask (resuscitator) – neonatal· paediatric and adult sizes | 6/12 (50%) | 6/12 (50%) | 0/12 (0%) | 0/12 (0%) |
| Compression bandages | 3/12 (25%) | 8/12 (66.6%) | 1/12 (8.3%) | 0/12 (0%) |
| Plasters or tape | 12/12 (100%) | 0/12 (0%) | 0/12 (0%) | 0/12 (0%) |
| Gauze | 11/12 (91.6%) | 1/12 (8.3%) | 0/12 (0%) | 0/12 (0%) |
| Intravenous cannulas of pediatric and adult sizes | 12/12 (100%) | 0/12 (0%) | 0/12 (0%) | 0/12 (0%) |
| Intravenous giving sets | 10/12 (83.3%) | 1/12 (8.3%) | 1/12 (8.3%) | 0/12 (0%) |
| Skin disinfectant for cannulation | 10/12 (83.3%) | 2/12(16.6%) | 0/12 (0%) | 0/12 (0%) |
| Syringes | 12/12 (100%) | 0/12 (0%) | 0/12 (0%) | 0/12 (0%) |
| Nutrition | 5/12 (41.6%) | 6/12 (50%) | 1/12 (8.3%) | 0/12 (0%) |
| Nasogastric tubes | 11/12 (91.6%) | 1/12 (8.3%) | 0/12 (0%) | 0/12 (0%) |
| Lubricant for nasogastric tube insertion | 6/12 (50%) | 6/12 (50%) | 0/12 (0%) | 0/12 (0%) |
| Intramuscular needles | 7/12 (58.3%) | 3/12 (25%) | 1/12 (8.3%) | 1/12 (8.3%) |
| Intraosseous cannulas of different sizes | 0/12 (0%) | 8/12 (66.6%) | 4/12 (33.3%) | 0/12 (0%) |
| Blankets | 7/12 (58.3%) | 5/12 (41.6%) | 0/12 (0%) | 0/12 (0%) |
| Facemasks for Infection Prevention and Control | 10/12 (83.3%) | 2/12(16.6%) | 0/12 (0%) | 0/12 (0%) |
| Aprons or gowns | 3/12 (25%) | 9/12 (75%) | 0/12 (0%) | 0/12 (0%) |
| Charts/notes for documentation | 11/12 (91.6%) | 1/12 (8.3%) | 0/12 (0%) | 0/12 (0%) |
| Pens | 10/12 (83.3%) | 1/12 (8.3%) | 1/12 (8.3%) | 0/12 (0%) |
| **Drugs** |  |  |  |  |
| Oral rehydration solution | 10/12 (83.3%) | 2/12(16.6%) | 0/12 (0%) | 0/12 (0%) |
| Intravenous crystalloid fluids | 11/12 (91.6%) | 1/12 (8.3%) | 0/12 (0%) | 0/12 (0%) |
| Intravenous dextrose fluid | 9/12 (75%) | 3/12 (25%) | 0/12 (0%) | 0/12 (0%) |
| Oxytocin | 7/12 (58.3%) | 5/12 (41.6%) | 0/12 (0%) | 0/12 (0%) |
| Adrenaline | 8/12 (66.6%) | 4/12 (33.3%) | 0/12 (0%) | 0/12 (0%) |
| Appropriate antibiotics | 4/12 (33.3%) | 8/12 (66.6%) | 0/12 (0%) | 0/12 (0%) |
| Diazepam | 9/12 (75%) | 3/12 (25%) | 0/12 (0%) | 0/12 (0%) |
| Magnesium sulphate | 8/12 (66.6%) | 4/12 (33.3%) | 0/12 (0%) | 0/12 (0%) |
| Paracetamol | 10/12 (83.3%) | 2/12(16.6%) | 0/12 (0%) | 0/12 (0%) |
| Local anaesthetic | 8/12 (66.6%) | 4/12 (33.3%) | 0/12 (0%) | 0/12 (0%) |
| **Human resources** |  |  |  |  |
| Health workers (e.g. nurses) with the ability to identify critical illness 24h/day | 11/12 (91.6%) | 1/12 (8.3%) | 0/12 (0%) | 0/12 (0%) |
| Health workers with the (e.g. nurses) ability to care for critically ill patients 24hrs/day | 8/12 (66.6%) | 4/12 (33.3%) | 0/12 (0%) | 0/12 (0%) |
| Senior health worker (e.g. doctor) who can be called to assist with the care of critically ill patients 24hrs/day | 10/12 (83.3%) | 2/12(16.6%) | 0/12 (0%) | 0/12 (0%) |
| **Training** |  |  |  |  |
| The health workers are trained in the identification of critical illness | 5/12 (41.6%) | 7/12 (58.3%) | 0/12 (0%) | 0/12 (0%) |
| The health workers are trained in the care of critically ill patients | 6/12 (50%) | 6/12 (50%) | 0/12 (0%) | 0/12 (0%) |
| **Routines** |  |  |  |  |
| The hospital has well defined routines for the identification of critical illness | 6/12 (50%) | 6/12 (50%) | 0/12 (0%) | 0/12 (0%) |
| The hospital has well defined routines for managing critically ill patients | 6/12 (50%) | 6/12 (50%) | 0/12 (0%) | 0/12 (0%) |
| There is a routine for the provision of EECC without taking into account patients’ ability to pay | 5/12 (41.6%) | 7/12 (58.3%) | 0/12 (0%) | 0/12 (0%) |
| There are routines for who and how to call to seek senior help 24hrs/day, 7 days/week | 12 (91.6%) | 1/12 (8.3%) | 0/12 (0%) | 0/12 (0%) |
| There are routines for integrating EECC with other care including the definitive care of the underlying condition (e.g. use of condition-specific guidelines) | 7/12 (58.3%) | 5/12 (41.6%) | 0/12 (0%) | 0/12 (0%) |
| **Guidelines** |  |  |  |  |
| There are written guidelines for the identification of critical illness | 4/12 (33.3%) | 5/12 (41.6%) | 3/12 (25%) | 0/12 (0%) |
| There are written guidelines for the essential care of critically ill patients | 3/12 (25%) | 6/12 (50%) | 3/12 (25%) | 0/12 (0%) |
| **Infrastructure** |  |  |  |  |
| Designated triage area (area for the identification of critical illness) in the Out Patient Department or Emergency Unit (area of the hospital where patients arrive) | 12/(100%) | 0/12 (0%) | 0/12 (0%) | 0/12 (0%) |
| Running water | 4/12 (33.3%) | 7/12 (58.3%) | 1/12 (8.3%) | 0/12 (0%) |
| Designated space for the care of critically ill patients (e.g. a bay· ward· high care unit) | 8/12 (66.6%) | 3/12 (25%) | 1/12 (8.3%) | 0/12 (0%) |
| Areas for separating and managing patients with a suspected or confirmed contagious disease from those without | 6/12 (50%) | 6/12 (50%) | 0/12 (0%) | 0/12 (0%) |

**Supplementary Table 2**: Thematic analysis of barriers to the implementation of EECC from healthcare providers’ perspectives

| **Theme** | **Subthemes** |
| --- | --- |
| Challenges in Early Identification | Staff shortage, poor monitoring, delayed recognition, communication gaps |
| Resource and Readiness Constraints | Equipment shortages, medication shortages, procurement barriers, infrastructure limitations |
| Knowledge and Competency Gaps | Limited training, skill decay, lack of simulation, weak competency assessment |
| Attitudes and Perceptions | Preference for advanced care, referral mentality, EECC undervaluation |
| Organizational Barriers | Staffing policies, budget constraints, EMR limitations |
| Motivation and Engagement | Incentives, recognition, burnout, ownership |
| Barriers to Timely EECC Delivery | Workload, resource delays, financial barriers, off-hour challenges |
